# biobank.cy: The Biobank of Cyprus past, present and future

**DOI:** 10.1101/2024.01.16.24301343

**Authors:** Eleni M. Loizidou, Maria Kyratzi, Maria A. Tsiarli, Andrea C. Kakouri, Georgia Charalambidou, Stella Antoniou, Stylianos Pieri, Panagiota Veloudi, Michaela Th. Mayrhofer, Andrea Wutte, Lukasz Kozera, Jens Habermann, Heimo Muller, Kurt Zatloukal, Karine Sargsyan, Alexandros Michaelides, Maria Papaioannou, Christos Schizas, Apostolos Malatras, Gregory Papagregoriou, Constantinos Deltas

**Author notes:** Corresponding author: Prof. Constantinos Deltas, University of Cyprus, School of Medicine 1, University Avenue, 2109 Nicosia.

## Abstract

**Background:** The Cyprus Biobank collects biosamples, medical and lifestyle information with the aim of reaching 16,500 Cypriots aged ≥18-years, by year 2027, as part of a multitasked EU funded project. Volunteers are both from the general population and from disease cohorts of focused research projects, who amongst others will contribute to canvas the architecture of the Cyprus human genome and study the healthy and morbid anatomy of Cypriots. The Cyprus Biobank is a research infrastructure pillar of the biobank.cy Center of Excellence in Biobanking and Biomedical Research.

**Methods:** Within 3-years (November 2019-October 2022), 1348 participants of the general population who represent a subset of the Cyprus Biobank recruited individuals, were enrolled in the pilot study. The study did not include individuals from separate disease-specific cohorts. Extensive information was collected from each participant, including biochemistry, complete blood count, physiological, anthropometric, socio-demographic, diet, and lifestyle characteristics. Prevalent health conditions along with medication use and family history were recorded, including 58 biomarkers based on blood and urine samples. With a systematic recruitment campaign, the Biobank is continuously increasing the number of individuals in the general population cohort and is developing separate disease cohorts of the Cypriot population.

**Results:** The pilot study enrolled 579 men and 769 women, aged between 18-85 years (median 48-years). The enrollment takes 40 minutes on average, including the collection of biological samples and phenotypic information. More than half (n=733, 55%) of the participants are educated to college level or above. Statistically significant differences were found between men and women regarding their education level (p<0.001), marital status (p=0.01) and employment status (p<0.001) but not their age (p=0.29). The most prevalent medical conditions recorded are hypertension (17.2%), osteoporosis (6.9%) and diabetes (6.0%).

**Conclusions:** The Cyprus biobanking pilot study has successfully collected extensive baseline information from enrolled participants. The Biobank will comprise a rich data resource used to examine the major risk factors leading to public health burdens and develop strategies for disease prevention.

## Introduction

Cyprus is the third largest island in the Mediterranean Sea and has been at the crossroad of many civilizations over the centuries. Its prestigious strategic location within the southeast corner of the Mediterranean basin, bridging the Middle East and Europe, always attracted foreign rulers that sought to control the area and thereby extend the trade pathways. The island of Cyprus was first inhabited during the aceramic neolithic period by people from the Middle East and Asia Minor, more than 12,000 years ago, according to archeological evidence and genetic phylogeographic investigations[1]. Over the years it was influenced, or was occupied by, several rulers (Table 1), the most recent occupations of which were the Ottoman Empire between the years of 1571-1878, after which it was taken over by the British, 1878-1960. After a liberation struggle, 1955-1959, the Republic of Cyprus was first established as an independent country in 1960, at which time it joined membership to the United Nations. In 2004, Cyprus joined the European Union and in 2008 adopted the Euro currency. The island’s recent and mainly older tumultuous history, including major and minor migration waves, has had its contribution to the Cypriot gene pool, as evidenced by several genetic studies performed over the past 30 years [2].

**Table 1.**
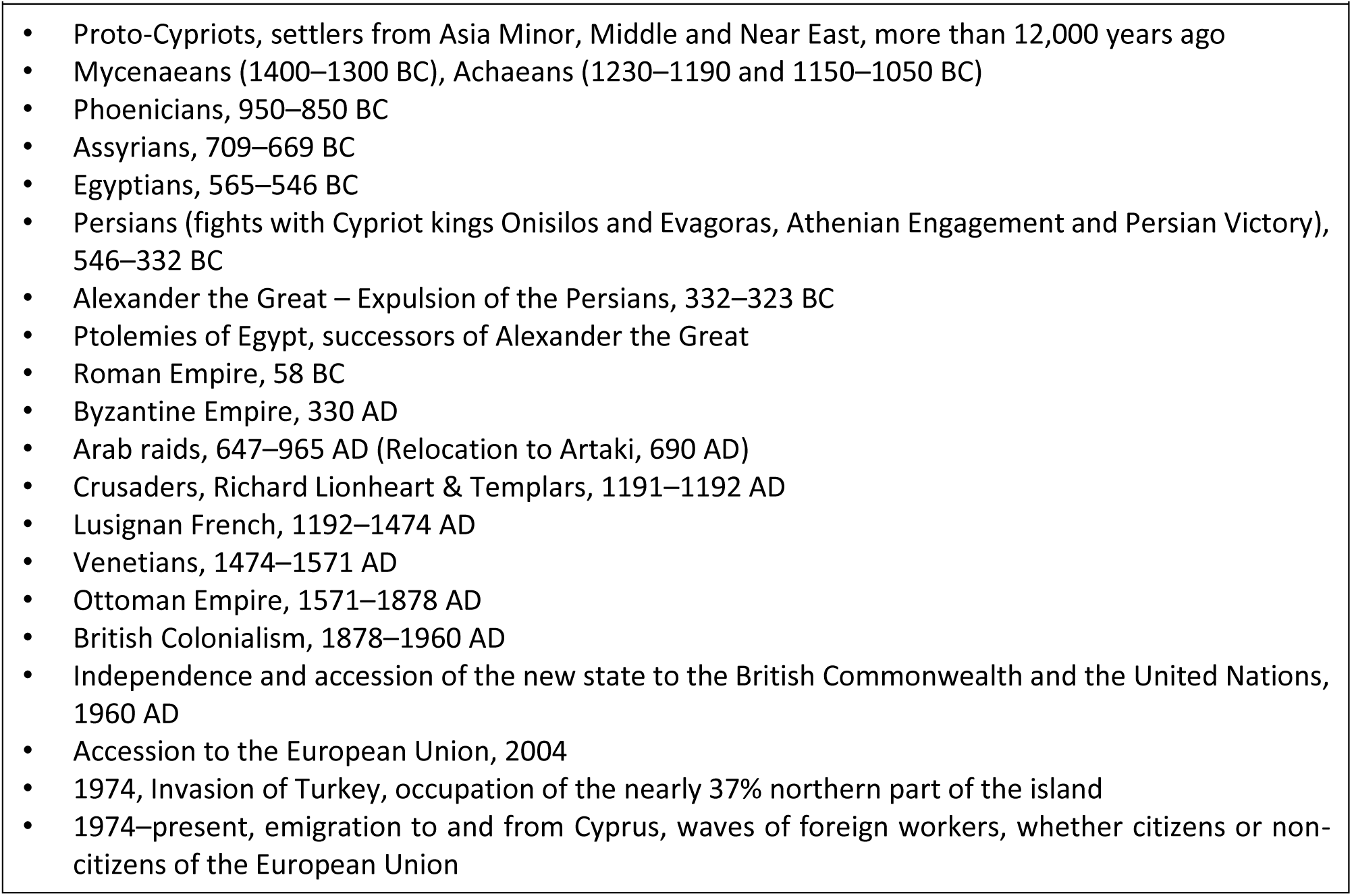
Brief report of visitors-conquerors and key events in the history of Cyprus. The Cypriot population adopted the Greek culture, language and religion since the Mycenaean Greeks reached the island, nearly 3,500 years ago. Several migration waves shaped the population gene pool. Three minorities were formed, Armenians and Maronites who arrived in several waves after the 6^th^ and 7^th^ century AD, respectively, while the Latins represent the remnants of the catholic Francs during the reign of the Lusignan, 12^th^-15^th^ century ([2]).

In the past 50 years, the Cypriot population has experienced major economic and lifestyle changes, due to major historical events and the economic crisis that has impacted especially the Eastern European countries in the past 10 years. Since the Turkish invasion in 1974, the population of Cyprus in the European non-occupied part of the island has increased from 648,585 to 923,272 individuals [3], not necessarily of Greek or Turkish origin. According to a 2021 census, the population of Cyprus altogether was estimated at 74% Greek Cypriots, including three small religious minorities of the Maronites, Armenians, and Latins, as well as Turkish Cypriots, altogether comprising about 1% of the population[3]. The population living in the northern part of the island under Turkish control, is estimated to be about 230,000. Despite the recent COVID-19 pandemic, life expectancy has only slightly changed between 2015 and 2021 (81.7 vs 81.2 years on average, respectively).

Chronic non-communicable diseases (NCDs), such as cardiovascular disease (CVD), diabetes, thyroid disorders, and cancer, pose a substantial health burden, significantly impacting morbidity and mortality rates. According to a 2018 report by the World Health Organization (WHO), NCDs accounted for over 90% of morbidity in Cyprus [4]. This prevalence of NCDs can be attributed to factors including lifestyle, dietary habits, and tobacco use. Also, genetic factors and gene-environment interactions contribute significantly to NCD development. Regional variation in NCDs has also been reported, suggesting the involvement of non-genetic factors that have yet to be identified. However, the presence of potential limitations in sample collection, such as the lack of representative samples and under-representation of specific regions may contribute to the fragmented understanding of NCD landscape in Cyprus [4]. From available information, cardiovascular disease is the primary cause of death amongst Cypriots, while the incidence of renal replacement therapy is 284 per million population, the highest in Europe [5].

Although most Cypriots declare a healthy status, two out of five suffer from a chronic disease, with CVD, cancer, and diabetes being the most prevalent conditions [6]. While CVD rates have been steadily decreasing since 2015, cancer rates remain at the same levels, albeit lower compared to the rest of the EU. On the contrary, tobacco use rates in Cyprus are amongst the highest in the EU, and together with unhealthy dietary habits, they comprise two of the most frequent mortality risk factors in Cyprus (19% and 14%, respectively) [6]. Additionally, while obesity rates in Cyprus are comparable to the average rates of other EU countries, childhood obesity is rapidly increasing and poses a significant public health issue for Cyprus.

To gain insights into the etiology and risk factors associated with NCDs and address limitations including the sampling of data from a diverse and representative population, the establishment of a Cypriot Biobank was imperative. While some causes of these diseases are already known based on studies conducted in other populations and regions [7, 8] current knowledge is insufficient to explain the substantial geographic differences in disease rates observed globally and within Cyprus itself [9]. The development of a Cypriot Biobank will enable large-scale studies of both genetic and environmental contributions, as well as their interactions, in the development of chronic diseases. Furthermore, it will address the limitations of previous studies, including inadequate sample sizes, lack of blood and/or urine samples, inconsistencies in data collection and processing, limited data collection on risk exposures (e.g. lifestyle, diet, disease comorbidities) and outcomes, and restricted study populations.

The biobank.cy Center of Excellence (CoE) in Biobanking and Biomedical Research was established in 2019 at the University of Cyprus (UCY), after acquiring funding through the Horizon 2020 Widening Participation/Teaming program (Grant Agreement no. 857122). The program is a partnership of the UCY with two Advanced Partners, the European Biobanking consortium BBMRI-ERIC [10] and the Medical University of Graz [11], and a local Partner, the medium-sized enterprise TALOS Rtd LTD [12]. Through this partnership, biobank.cy has successfully received funding to advance the country’s medical research infrastructure to match European standards, with two major objectives: a) Upgrade the first Biobank of the country and complete the Cyprus Human Genome Project; b) build the capacity and upgrade its research infrastructure with the aim to prepare Cyprus for the next generation of biomedical research. For the history, biobank.cy is the result of upgrading the Molecular Medicine Research Center (MMRC) which was first established in 2010 with another strategic funding of the Regional Development Fund of the European Union and the Republic of Cyprus through the Cyprus Research Promotion Foundation, as part of the Smart Specialization Strategy of the country. In its current status, the Biobank is one of five Pillars of the biobank.cy CoE, the other four being, the MMRC as the research arm of the CoE, and the Molecular Diagnostics, the Education and Innovation pillars.

Towards the public health benefit and to support the development of the Cyprus Human Genome Project, the Government of Cyprus, and the University of Cyprus co-funded biobank.cy. As a first milestone, up to 16,500 permanent Cypriot residents will be invited to partake in the biobank cohort, voluntarily, until 2027. Data collection and storage includes baseline clinical, biochemical, and behavioral traits as well as blood and urine samples.

We herein provide comprehensive information on the operation of the first Biobank in Cyprus, sharing data on the demographics and health archives of participants.

## Methods

### Biobank setting

The biobank.cy, as an EU-funded CoE in Research and Innovation, hosts and maintains the Cyprus Biobank, which is not based in a hospital setting but enrolls volunteers who respond to open invitations while also collaborating with public and private clinics and ad hoc volunteer enrolment centers around the island. Operation of the Biobank in Cyprus is not regulated by a specialized law, although a legislation proposal regulating biobanks creation and operation is under consideration. There are national and EU laws regarding regulating the use of biological samples for research [13], the law providing for the protection of natural persons with regard to the processing of personal data and for the free movement of such data[14], the law for the establishment of the Cyprus National Bioethics Committee (CNBC)[15], and the EU GDPR 2016/679 law for the General Data Protection Regulation, which plays a significant role in emphasizing data protection principles, as the Biobank is in full compliance to its provisions[16]. These laws collectively ensure data protection, ethical standards, and informed consent in Biobank operations.

Approval for the operation of this first Biobank was first obtained by the CNBC in November 2011 (approval code: EEBK/ΕΠ/2011/04). The approved consent form allows for broad use of the data and material in Cyprus and abroad provided that any future research project secures its own separate ethics approval (English version of consent form is Supplementary Material_1).

### Data collection and storage

The Biobank gathers information through a structured and comprehensive questionnaire, clinical phenotyping, and biological samples collected from Cypriot residents of the general population, aged ≥ 18 years old. Population data of the pilot study are collected and stored using REDCap® [17, 18] and OpenSpecimen® [19] tools respectively, to enable data security and tracking as well as to eliminate collection bias. Specifically, REDCap® (Research Electronic Data Capture) is a secure, web-based application designed for data collection and management in research studies, used to amass the following measurements: personal and family history as well as clinical (complete blood count, biochemical) and biometric (blood pressure, height, weight) (Supplementary Material_2). OpenSpecimen® is used as the Biobank’s Information Management System, by organizing information related to the collection, processing, storage, and distribution of samples.

### Participant/volunteer enrolment

Since the very first enrollment on the 1^st^ of November 2019, participant awareness and engagement in the project were achieved through media and the press (television, radio, social media and newspapers, leaflets respectively) as well as social events which are organized by the Biobank personnel. Additionally, friends and family referrals were frequent throughout the 3-year period. Interested individuals book an appointment either through the biobank.cy website [20] or directly through a phone call. At baseline, participants are invited to the central collection point of the Biobank facility at the Shacolas Educational Center for Clinical Medicine, or elsewhere during outreach activities, where they undergo a 3-stage interview (25 min on average) and physical and clinical measurements (15 min on average) (Figure 1; Table 2). The Biobank participation process commences with trained research nurses providing an initial briefing to the participants. During this session, participants are fully informed about the research project’s purpose, their rights, and the procedures for participation and withdrawal, while they may ask questions about the project, prior to signing a broad consent form. The consent form is approved by the Cyprus National Bioethics Committee (approval code: EEBK/ΕΠ/2020/04), ensuring compliance with ethical requirements. A copy of the signed consent form is given to the participant.

**Figure 1:**
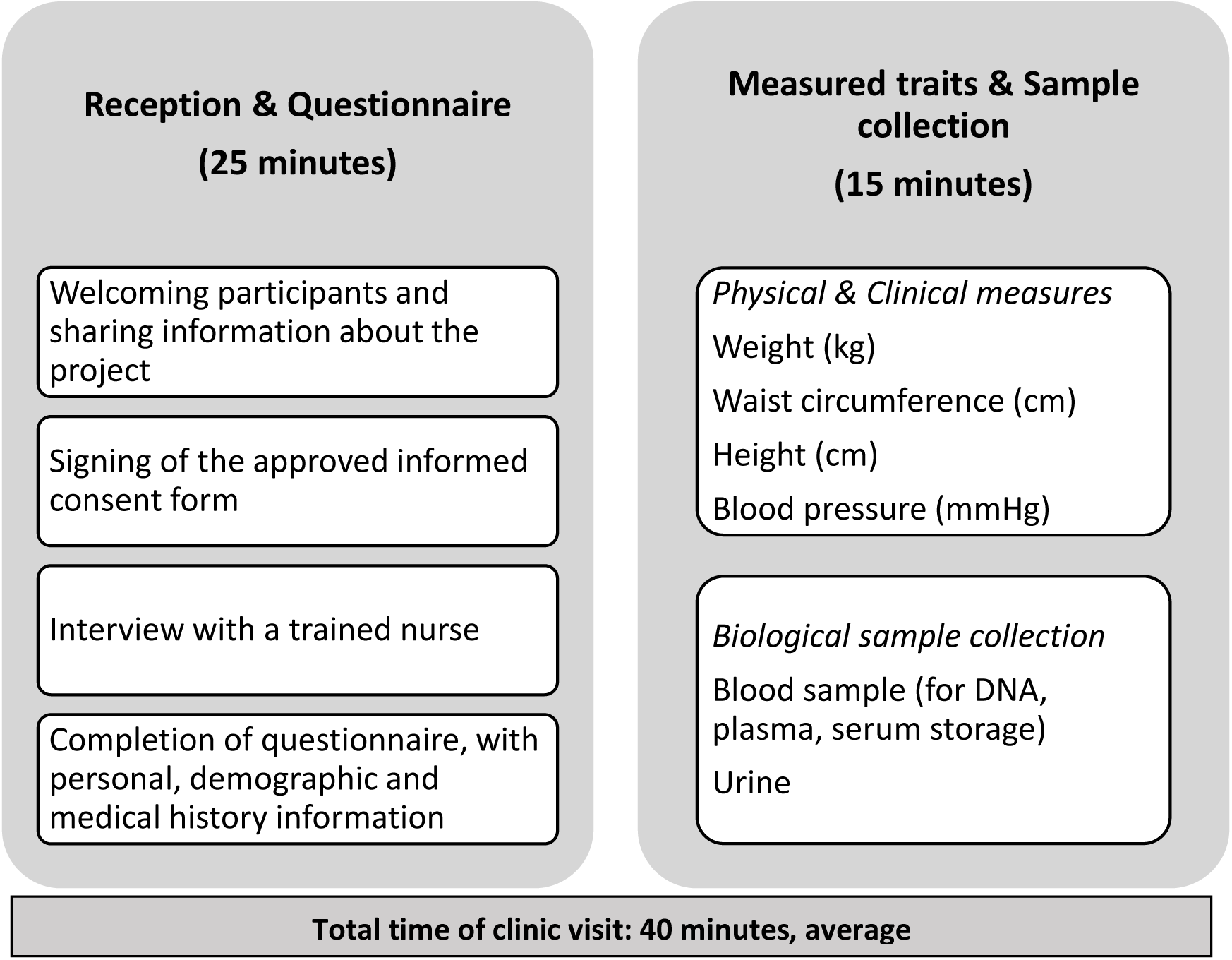
Baseline visit summary.

**Table 2:** Instruments used at the Biobank for data measurements.

**Figure 1: Baseline visit summary**
| **Table 2: Instruments used at the Biobank for data measurements** |  |
| --- | --- |
| **Measurement category** | **Instrument** |
| **Questionnaires** |  |
| Health & Lifestyle | Questions on socio-demographic factors, medical conditions, medication use, family history, smoking behavior, alcohol consumption, physical activity levels and sleeping patterns |
| Nurse Interview | Additional questions on QoL assessment (smoking, alcohol consumption, and physical activity frequency), family status, doctor-diagnosed conditions, education, employment status, and dietary patterns |
| **Physical and clinical measurements** |  |
| Anthropometric | Height, weight, and waist circumference measurements* |
| Physiological | Blood pressure using a digital sphygmomanometer |
| **Biological samples** |  |
| Biological samples collected | Whole peripheral blood, urine |

\*Waist circumference measures available for 960 individuals

Upon completion of the informed consent process, participants are requested to fill out a questionnaire providing essential information about their personal and family medical history. This information is securely collected in dedicated REDCap® projects.

### Health and lifestyle questionnaire administered by a trained research nurse

In addition to data of personal character, the health and lifestyle section of the questionnaire includes questions on socio-demographic factors, including the origin of both parents, medical conditions along with medication used, family history and quality-of-life (QoL) questions, including smoking behavior, dietary habits, alcohol consumption, intensity of physical activity [21] and sleeping patterns [22], education level and employment status. As the questionnaire is part of the registration and clinic visit, a Biobank-trained research nurse is always available to answer any questions of the participant.

Importantly, questions on dietary assessment are directly linked to the Mediterranean score (MedDietScore) [23] which indicates the level of the participants’ adherence to the traditional Mediterranean diet. This specific type of diet has been widely used for the past ∼20 years as it has been proven to significantly reduce total mortality [24], thus having a great impact on clinical and public health. More specifically, a higher MedDiet score showed strong evidence against incidents of cardiovascular disease, diabetes, hypertension, obesity, and Alzheimer’s disease among others [25]. Participants are specifically asked to report the consumption frequency of whole grain food (bread, pasta, rice, etc), potatoes, fruit and juice, vegetables and salads, legumes, fish and soup, red meat and its products/derivatives, poultry, full-fat dairy, and olive oil.

### Physical-clinical measurements and biological material collected

At baseline, anthropometric and physiological traits were measured for each Biobank participant along with blood and urine sample collection. Anthropometric and physiological traits comprised body weight/height and diastolic/systolic blood pressure respectively. Blood pressure was measured once, in a seated position. A blood pressure measurement was repeated if the first measurement returned a value of >160mmHg for systolic blood pressure or <80 mmHg for diastolic blood pressure.

At the baseline enrolment visit, the participants provided peripheral blood for the isolation of DNA, plasma, and serum, which are stored at −80°C. Urine was also collected and aliquots were stored likewise. Specifically, we collected: 1×10ml EDTA for DNA extraction, 1×4ml EDTA for FBC, 1×4ml EDTA for plasma, 1×8.5ml serum, 1×3.5ml serum (prompt test), and 1×100ml urine. Aliquoted and stored are 4 aliquots of 50μl DNA at 50ng/μl, plus a concentrated DNA stock, 4 aliquots of 2ml serum, 2 aliquots of 1ml plasma, and 5 aliquots of 2ml urine.

Fifty-eight clinical biomarkers are measured by a private diagnostic provider using a proportion of the stored blood (Table 3). DNA concentration and quality are tested by spectrophotometric measurements and calculation of the ratio at OD 260/280 and OD at 260/230. Urine is stored after a 5-minute centrifugation at 1000g at room temperature. The quality of frozen serum samples is assured in random retesting of selected biochemical parameters.The Biobank employs a data curator who constantly and systematically tests for quality assurance and consistency of data input to minimize errors and avoid bias. A comprehensive timeline of biobanking and data/sample retrieval, with brief information on Standard Operating Procedures (SOPs) and quality assurance measures applied throughout the procedure is provided in Figure 2. The detailed standard operating procedures used at the Biobank are available on request.

**Figure 2:**
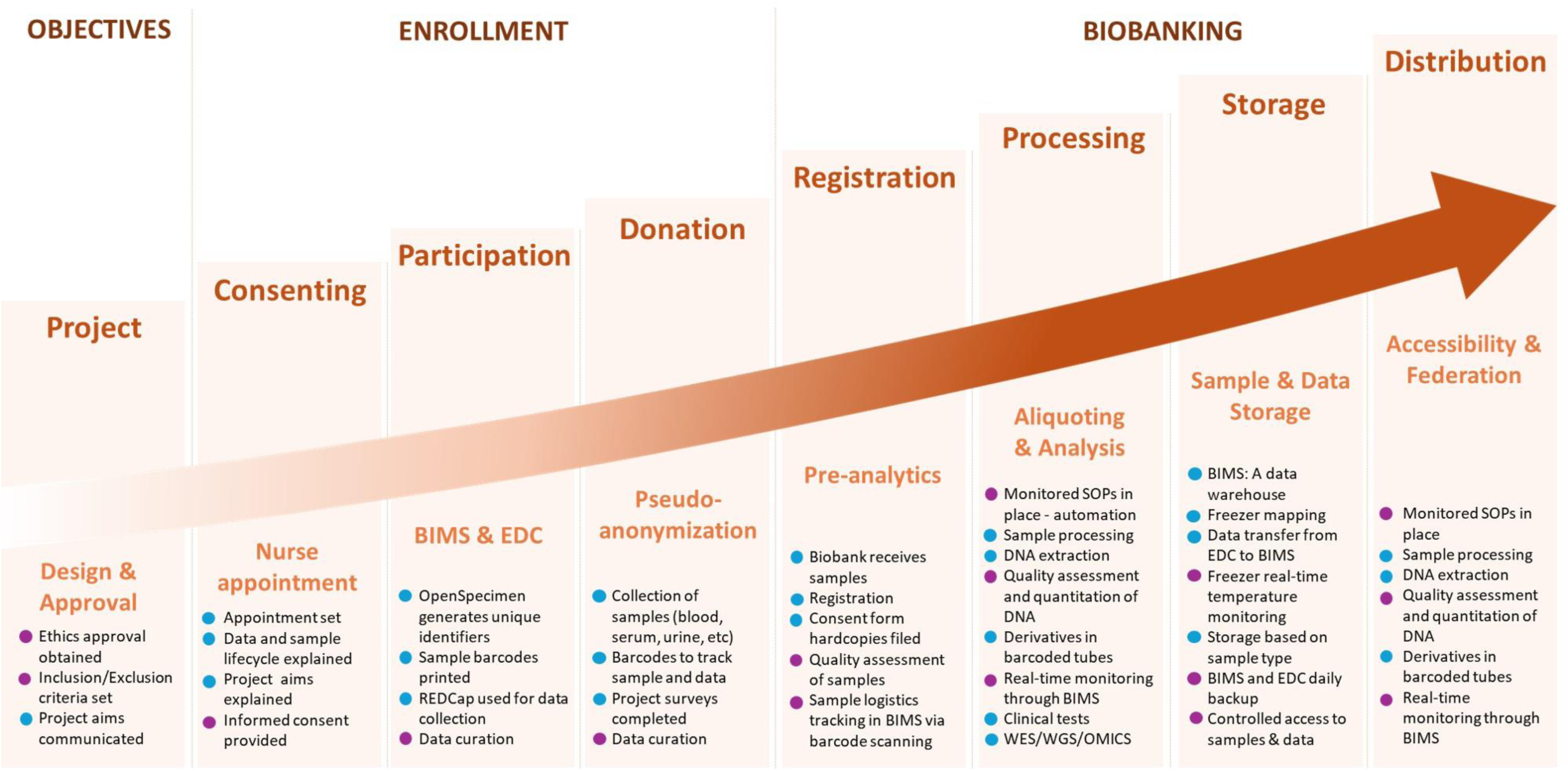
Timeline of biobanking and data/sample retrieval, with brief information on SOPs and quality assurance measures applied throughout the procedure. The purple dots represent the quality assurance checkpoints and the blue dots represent the activities based on monitored SOPs.

**Table 3:** Clinical biomarkers measured at biobank.cy (N = 58)

| **Table 3: Clinical biomarkers measured at biobank.cy (N = 58)** |  |  |
| --- | --- | --- |
| **Biomarker group** | **Measure** | **Units** |
| **Blood biochemistry** |  |  |
| Lipid profile | Total cholesterol | mg/dl |
|  | Low-density lipoprotein (LDL) | mg/dl |
|  | High-density lipoprotein (HDL) | mg/dL |
|  | Triglycerides | mg/dL |
| Diabetes-related tests | Glucose | mg/dL |
|  | Glycated Haemoglobin A1c % | % |
| Bone and joint markers | Uric acid | mg/dL |
| **Complete Blood Count** |  |  |
| White blood cells | White blood cells | $\times 10^3/\mu\text{L}$ |
| Differential white cell count | Lymphocytes | $\times 10^3/\mu\text{L}$ |
|  | Lymphocytes % | % |
| | Eosonophils | $\times 10^3/\mu\text{L}$ |
|  | Eosonophils % | % |
| | Monocytes | $\times 10^3/\mu\text{L}$ |
|  | Monocytes % | % |
| | Basophils | $\times 10^3/\mu\text{L}$ |
|  | Basophils % | % |
| | Neutrophils | $\times 10^3/\mu\text{L}$ |
|  | Neutrophils % | % |
| | LUC | $\times 10^3/\mu\text{L}$ |
|  | LUC % | % |
| Red blood cells | Haemoglobin | gr/dL |
| | Red blood cells | $\times 10^6/\mu\text{L}$ |
|  | Haematocrit | % |
|  | Mean Corpuscular Hemoglobin (MCH) | pg |
|  | Mean Corpuscular Hemoglobin Concentration (MCHC) | g/dL |
|  | Mean Corpuscular Volume (MCV) | femtoliters (fL) |
|  | Red Cell Distribution Width coefficient of variation (RDW CV) | % |
|  | Haemoglobin Distribution Width (HDW) | % |
| Platelets | Platelets | $10^3/\text{mm}^3$ |
|  | Platelet Distribution Width (PDW) | % |
|  | Mean Platelet Volume (MPV) | femtoliters (fL) |
|  | Plateletcrit (PCT) | % |
| **Urine Biochemistry by dipstick, and Microscopy** |  |  |
| Biochemistry | Protein | mg/dL |
|  | Glucose | mg/dL |
|  | Ketones | Negative/Positive |
|  | Bilirubin | Negative/Positive |
|  | Urobilinogen | mg/dL |
|  | Nitrates | Negative/Positive |
| | Erythrocytes/Haemoglobin | Ery/$\mu\text{L}$ |
| | Leucocytes | Leuco/$\mu\text{L}$ |
|  | Reaction (pH) | pH: 5-9 |
|  | Specific gravity | 1,000 - 1030 |
| Macroscopy | Appearance |  |
|  | Colour |  |
| Microscopy | WBC (/hpf) |  |
|  | RBC (/hpf) |  |
|  | Epithelial cells |  |
|  | Casts |  |
|  | Casts Hyaline |  |
|  | Cast Granular |  |
|  | Crystals |  |
|  | Microorganisms |  |
|  | Yeasts |  |

| Urine Biochemistry |  |  |
| --- | --- | --- |
| Microalbumin | Albumin | mg/L |
|  | Microalbumin | mg/dl |
|  | Urine Albumin/Creatinine ratio | mg/gr Creat |
| Protein/Creatinine Urine Ratio | Proteins | mg/dl |
|  | Creatinine | mg/dl |
|  | Urine Protein/Creatinine ratio | mg/gr Creat |

## Results

### Sample size

We performed a power calculation using the "pwr" package in R software to estimate the required sample size for the Cypriot Biobank, based on a population of 923,372 individuals. The calculation was conducted using a 5% significance level and we adjusted for an expected response rate of 80%, which resulted in a required sample size of 480 individuals. In determining the sample size, we combined the power calculation with practical considerations. We accounted for factors such as the number of research nurses that would be hired, the expected number of participants per day after discussion with local clinicians and healthcare providers and the capacity of our facilities. Furthermore, we reviewed the approaches used by established biobanks to determine appropriate sample sizes. Burton et al[26], emphasize that biobanks intended for human genome epidemiology studies should consider using larger sample sizes to address potential heterogeneity in disease risk, enhance the precision of estimates, and improve sensitivity and specificity in diagnostic testing. Using this integrated approach of the theoretical sample size calculation, educated guesses and practical considerations, we estimated that a sample size of 16,500 individuals would be sufficient for the Biobank to be scientifically robust and operationally feasible. The pilot study enrolled 1348 participants not included in any disease-specific cohort of the Biobank, with a geographical distribution based on the father’s origin (Figure 3).

**Figure 3:**
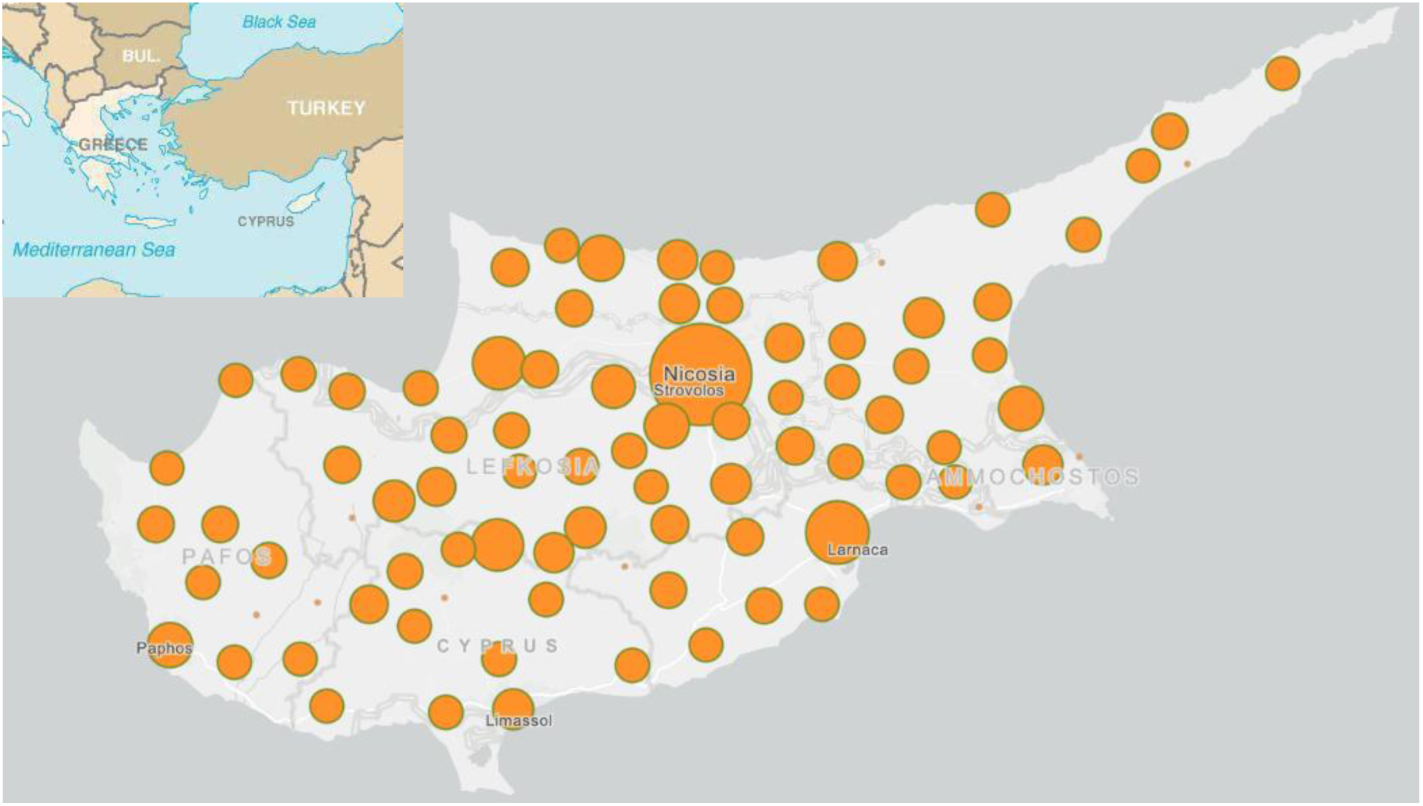
Geographical distribution of the 1348 participants of the pilot study, based on the origin of the father. The circle diameter reflects the density of the donors’ enrolment in the corresponding area.

### Socio-demographic characteristics

The participants of the Biobank pilot study comprised 579 men (43%) and 769 women (57%) spanning an age range from 18-85 years, with a median age of 48 years (SD = 13.9 years) (Figure 4). More than half of men (n = 323 (55.8%)) and women (n = 420 (54.6%)) were educated to a university degree level and above, while the most frequent employment status for both men and women was paid employee (n = 358 (62%) and n = 486 (63%) respectively) (Table 4). The most common status of the enrolled individuals was either single or married (n = 282 (20.9%) and n = 981 (72.8%) respectively).

**Figure 4:**
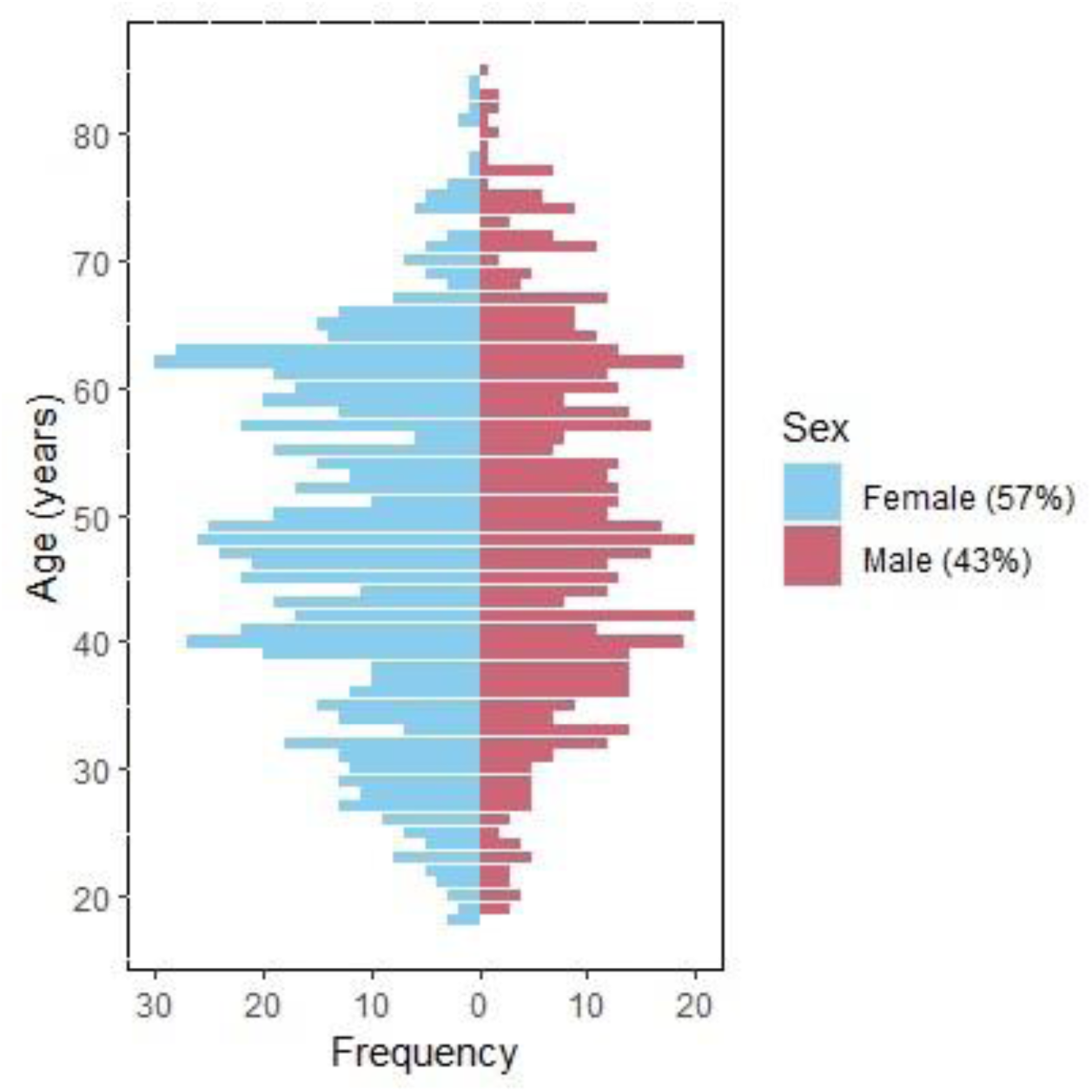
Age distribution of men and women enrolled in the Biobank pilot study

**Table 4:** Socio-demographic characteristics of the enrolled participants in the Biobank pilot study (November 2019 – October 2022). P-values for the comparison between sex groups were calculated using Chi-squared and Fisher’s exact tests.

| Characteristic | Men (%) | Women (%) | P-value |
| --- | --- | --- | --- |
|  | 579 (43) | 769 (57) |  |
| **Age group (years)** |  |  | 0.29 |
| < 25 | 22 (3.8) | 30 (3.9) |  |
| 25-34 | 65 (11.2) | 116 (15.1) |  |
| 35-44 | 135 (23.3) | 163 (21.2) |  |
| 45-54 | 141 (24.4) | 192 (25.0) |  |
| ≥ 55 | 216 (37.3) | 268 (34.9) |  |
| **Marital status** |  |  | 0.01 |
| Single | 128 (22.1) | 154 (20.0) |  |
| Married | 429 (74.1) | 552 (71.8) |  |
| Divorced | 18 (3.1) | 44 (5.7) |  |
| Widowed | 4 (0.7) | 18 (2.3) |  |
| Civil partnership | 0 (0) | 1 (0.1) |  |
| **Education** |  |  | < 0.001 |
| Lower than primary school | 1 (0.1) | 3 (0.4) |  |
| Primary school | 17 (2.9) | 34 (4.4) |  |
| Secondary school | 161 (27.8) | 167 (21.7) |  |
| Technical/professional school | 38 (6.6) | 21 (2.7) |  |
| College diploma (degree not equivalent to a BSc/BA) | 39 (6.7) | 124 (16.1) |  |
| Bachelor's degree | 200 (34.5) | 261 (33.9) |  |
| Master's degree | 91 (15.7) | 141 (18.3) |  |
| PhD degree | 32 (5.5) | 18 (2.3) |  |
| **Employment status** |  |  | < 0.001 |
| Paid employee | 358 (61.8) | 486 (63.2) |  |
| Self-employed | 50 (8.6) | 44 (5.7) |  |
| Housewife | 0 (0) | 41 (5.3) |  |
| Retired | 87 (15.0) | 76 (9.9) |  |
| Unemployed | 12 (2.1) | 27 (3.5) |  |
| Other | 33 (5.7) | 33 (4.3) |  |
| Not reported | 39 (6.7) | 62 (8.1) |  |

### Medical history

Based on the NHS and Heart UK guidelines (https://www.nhs.uk/conditions/high-cholesterol/cholesterol-levels/), nearly half of the participants had high total (n = 681 (50.5%)) and LDL (n = 630 (46.7%)) cholesterol levels. Additionally, 15.8% (n = 213) had low HDL levels and 19% (n = 257) had high triglyceride levels. Most of these participants did not fast prior to the blood sample analysis. Among those with abnormal levels, only 151 out of 681 (22.2%) with high total cholesterol, 150 out of 630 (23.8%) with high LDL cholesterol, 31 out of 213 (14.6%) with low HDL cholesterol, and 34 out of 257 (13.2%) with high triglyceride levels fasted before the tests. The results of our pilot study are in line with recent findings regarding the high prevalence of elevated blood lipid levels in the European population [27–29]￼.

Even though most participants self-reported as healthy, a varying percentage had underlying past or current conditions, with the most prevalent being hypertension (17.2%), osteoporosis (6.9%) and diabetes (6.0%) (Figure 5). These findings are in line with OECD’s research on the most common chronic conditions in Europe for 2022 (https://health.ec.europa.eu/system/files/2022-12/2022_healthatglance_rep_en_0.pdf). As useful as they are, they will prompt public policy decisions and enhance existing prevention programs. Additional conditions recorded include cardiovascular disease (myocardial infarction and stroke), rheumatoid arthritis, osteoarthritis, asthma, nephropathies, cancer and depression (Fig.4). Most individuals with a previously diagnosed condition were under medication use (Table 5).

**Figure 5:**
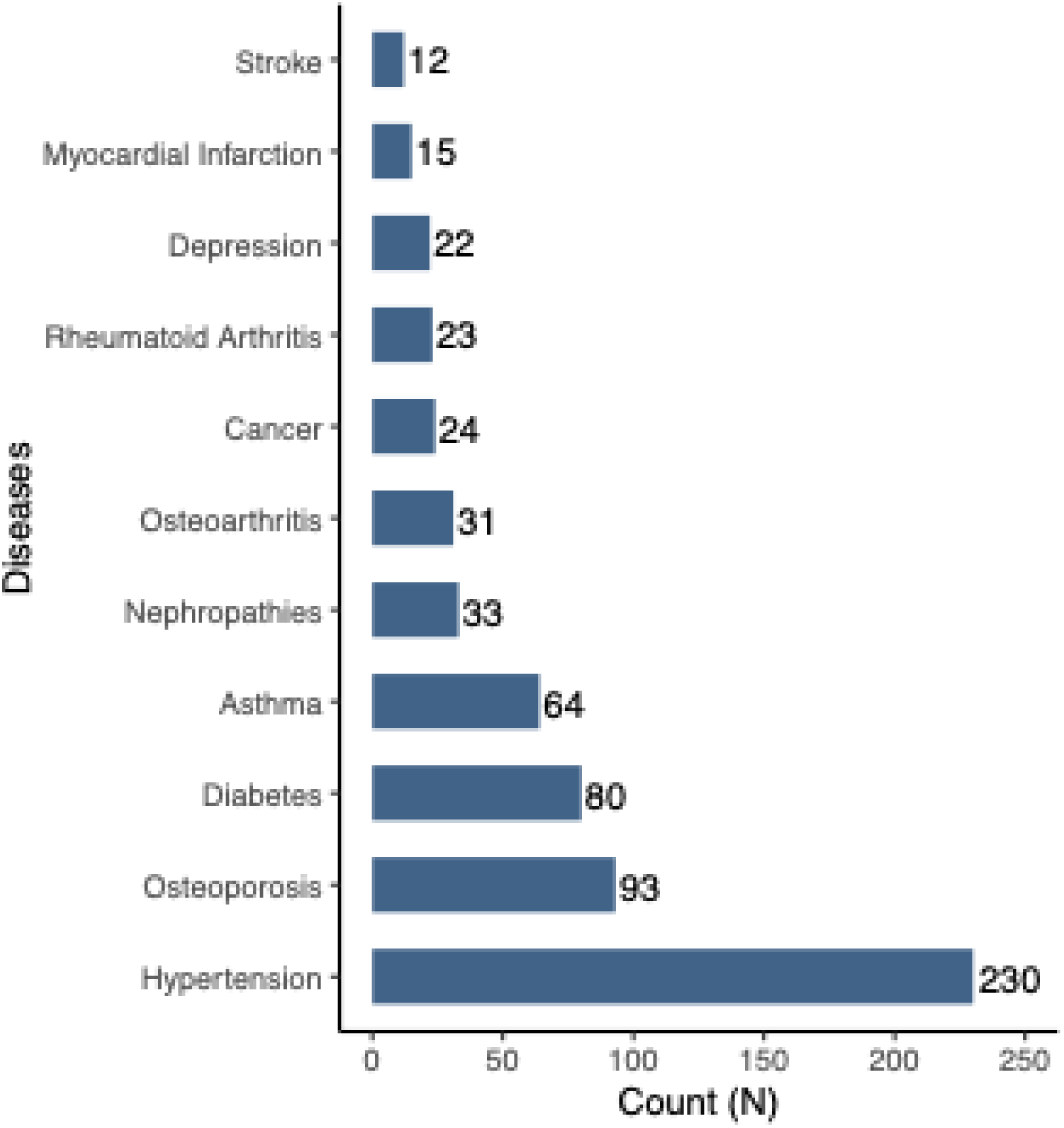
Diseases recorded among the participants of the pilot study

**Table 5:** Medical history and medication use (n = 1348). ^ǂ^The disease is self-reported according to the doctor diagnosis.

| **Table 5:** Medical history and medication use (n = 1348). †The disease is self-reported according to the doctor diagnosis. |  |  |  |  |  |  |  |  |  |
| --- | --- | --- | --- | --- | --- | --- | --- | --- | --- |
| **Disease†**<br>**(ICD10CM)** |  | **Sex** |  | **Age group N (%)\*** |  |  |  |  | **Medication use** |
|  | **N<sub>total</sub> (%)\*** | **N<sub>female</sub> (%)\*** | **N<sub>male</sub> (%)\*** | **<25** | **25-34** | **35-44** | **45-54** | **≥55** | **N (%)** |
| Hypertension (I10) | 230 (17.2) | 132 (57.4) | 98 (42.6) | 0 (0) | 1 (0.4) | 10 (4.3) | 47 (20.4) | 172 (74.8) | 219 (95.2) |
| Myocardial Infarction (I21.-) | 15 (1.1) | 13 (86.7) | 2 (13.3) | 0 (0) | 0 (0) | 1 (6.7) | 1 (6.7) | 13 (86.7) | 13 (86.7) |
| Stroke (I63.9) | 12 (0.9) | 6 (50.0) | 6 (50.0) | 0 (0) | 1 (8.3) | 2 (16.7) | 2 (16.7) | 7 (58.3) | 10 (83.3) |
| Diabetes (E11.8) | 80 (6.0) | 43 (53.8) | 37 (46.2) | 0 (0) | 0 (0) | 5 (6.2) | 23 (28.8) | 52 (65.0) | 78 (97.5) |
| Rheumatoid Arthritis (M06.9) | 23 (1.7) | 9 (39.1) | 14 (60.9) | 0 (0) | 1 (4.3) | 1 (4.3) | 2 (8.7) | 19 (82.6) | 21 (91.3) |
| Osteoarthritis (M19.9) | 31 (2.3) | 9 (29) | 22 (71) | 0 (0) | 0 (0) | 3 (9.7) | 4 (12.9) | 24 (77.4) | 29 (93.5) |
| Osteoporosis (M81.9) | 93 (6.9) | 13 (14) | 80 (86) | 0 (0) | 1 (1.1) | 3 (3.2) | 19 (20.4) | 70 (75.3) | 86 (92.5) |
| Asthma (J45.90) | 64 (4.8) | 29 (45.3) | 35 (54.7) | 3 (4.7) | 3 (4.7) | 19 (29.7) | 14 (21.9) | 25 (39.0) | 45 (70.3) |
| Nephropathies (N28.9) | 33 (2.5) | 20 (60.6) | 13 (39.4) | 2 (6.0) | 1 (3.0) | 9 (27.3) | 4 (12.1) | 17 (51.5) | 27 (81.8) |
| Cancer (C00-C97) | 24 (1.8) | 10 (41.7) | 14 (58.3) | 0 (0) | 1 (4.2) | 0 (0) | 3 (12.5) | 20 (83.3) | 7 (29.2) |
| Depression (F32.9) | 22 (1.6) | 3 (13.6) | 19 (86.4) | 0 (0) | 2 (9.0) | 7 (31.8) | 5 (22.7) | 8 (36.4) | 17 (77.3) |

\* $N_{\text{total}}(\%)$ is the number of participants with the condition out of the total number of participants ( $n = 1348$ ). $N_{\text{male}}(\%)$ and $N_{\text{female}}(\%)$ represent the number of males and females with the condition respectively, out of the total number of participants with the condition ( $N_{\text{total}}$ ). Age group $N(\%)$ represents the number of participants with a condition by age group. Medication use $N(\%)$ indicates the number of participants under disease-specific medication use out of the total number of participants with the condition ( $N_{\text{total}}$ ).

### Collaborations and perspectives

The availability of the Biobank infrastructure has fostered collaborations with groups at national and international Universities and Research Centers, including Europe and the USA. Sharing of data and material is based on bipartite partnerships, either serving as custodians to data/sample archiving and storage or more intimately involved in common research projects and grant writing. Signed Material Transfer Agreement/Data Transfer Agreement (MTA/DTA) are in place, usually using our own approved documents. State-of-the-art equipment, software and expertise in several fields such as biostatistics, epidemiology, bioinformatics and genetics/genomics, have resulted in several important collaborations for massive genetic analyses (Whole Exome Sequencing) of diverse disease cohorts (e.g. inherited kidney diseases, inherited cardiomyopathies, traumatic brain injury). Also, the CYPROME data, based on 1000 Cypriot whole exomes, that is now used to serve as the Cypriot reference genome, has already started being instrumental in interpreting results from research projects and diagnostic services. Further infrastructure development includes purchasing robotic systems for substantially increasing the results output and turnover, while another larger server is pending for storing the Big Data generated in-house or through outsourcing and collaborations. Finally, to maximize the use of samples and data, the Biobank personnel is also closely engaged in research performing tasks.

## Discussion

### The CY-Biobank

The establishment of the first Biobank in Cyprus, in 2011, and its enhancement in 2019 as part of the biobank.cy Center of Excellence, provides an opportunity for ground-breaking research both nationally and internationally, through a rich collection of high-quality phenotypic data that will unveil the causes of common and rare diseases affecting the Cypriot population. The data are retrieved from organized questionnaires, the collection of blood and urine samples, and detailed clinical measurements. The Biobank aims to reach a total sample size of 16,500 individuals by 2027 and intends to perform follow-up studies in a sub-group of volunteers in the long-term to record disease progression, based on criteria to be determined. After informed consent from the participants, the data is stored for research purposes with the goal to canvas the architecture of the Cyprus human genome. biobank.cy is equipped with clinic, laboratory and liquid sample storage facilities and offices for non-clinical research staff.

The Biobank was founded and launched as a national research infrastructure in 2011 with national and EU funding and was significantly enhanced in 2019 through a Horizon 2020/Widening/Teaming Program, just before the COVID-19 pandemic started. Despite the challenges and the relatively short period since its establishment, it already has a high impact on the health of the Cypriot population. During the pandemic, in addition to the original scope, biobank.cy collected samples from COVID-19 cases to study the production of circulating immunoglobulin class G (IgG) antibodies against SARS-CoV-2 in the serum of convalescent individuals [30]. Individuals who participated in the project received the results from their sample collection two weeks later, thus gaining the opportunity to discuss potential abnormal ranges with their personal doctors. The whole exome sequencing (WES) of 1500 convalescent participants in the project is pending. Finally, notwithstanding the challenge of not being hosted at a hospital, we managed to turn it into an opportunity by organizing outreach excursions to the periphery or at the premises of large enterprises and organizations, for enrolling volunteers who would not easily reach us in Nicosia. It is of note that owing to the small geography of the island, samples reach the Biobank within a maximum of two hours after completion of the enrolments, by 2:00 pm the latest. This unrestricted enrolment strategy will enable the design of epidemiological and other public health studies including the establishment of longitudinal cohorts based on the data collected.

According to the data collected in this pilot study, volunteers diagnosed with hypertension represent 17.2% of the general population, diabetics represent 6%, patients with osteoporosis represent 6.9%, and patients with asthma represent 4.8%. With the understanding that these were not targeted patient groups, we anticipate that with the systematic projected increase in participant recruitment, the Cypriot population will soon be represented in greater depth which will validate and enhance our conclusions, with its separate disease cohorts available for further research.

### Strategy and vision

#### Cyprus Human Genome Project

biobank.cy was formed with the vision of being the island’s central node for archiving high-quality biomaterial and associated clinical records, with state-of-art research infrastructure facilities utilized for precision medicine. Towards this goal, biobank.cy has launched the largest research study to ever run in Cyprus, with the precise aim of understanding and treating diseases by characterizing the Cypriot genome. Specifically, biobank.cy has initiated the Cyprus Human Genome Project (CHGP). This is the first in its kind of scale and prospect study in Cyprus, with the aim of implementing Whole Exome Sequencing (WES) of 1000 Cypriots at phase-1 and up to 5000 in the next 3 years. Also, Whole Genome Sequencing is a strategic objective. This data will constitute the blueprint for the first Cypriot reference genome (CYPROME). Concurrently, additional volunteers of the general population, including chronic disease patients, are selected for WES based on additional ongoing separately funded research projects.

The establishment of the Biobank was a landmark development that enabled the materialization of the CHGP as a significant milestone and a turning point for the health of the Cypriot population and precision medicine. The CHGP will provide unprecedented insights in terms of the pathogenic variant load and disease prevalence in the Cypriot population, while it will be of use to targeted epidemiological studies across all disciplines and non-communicable disease investigations. Furthermore, the CHGP will provide the backbone to address specific diseases on the island by documenting their prevalence and revealing putative environmental factors that affect them. Moreover, biobank.cy with its increasing potential as a central medical research hub, will provide an unsurpassable opportunity for Cypriot researchers and medical professionals to expand their collaborations nationally and internationally while it will offer the opportunity to Cypriot patients to have access to clinical trials and treatments not currently available in Cyprus.

Over the past 30+ years, genetic studies in the Cypriot population unraveled many interesting findings concerning mainly rare diseases, including kidney conditions, neurological conditions, endocrinological conditions and others [2]. Founder mutations were discovered for several diseases while a Cypriot consensus whole genome sequence (including the non-coding part) is still lacking. The CHGP is promising to identify more DNA variants of pathogenic potential and enable comparative studies in relation to the minor allele frequencies among the neighboring populations, thus leading to a better understanding of our genetic architecture.

### Imminent Objectives

With regards to banking itself, a major effort is to increase the number of participants to cover the population to a greater depth, thereby empowering relevant epidemiological and genetics studies on all pathologies and enabling twin and population screening studies. To achieve our strategic objective of enrolling 16,500 participants by 2026 (now extended to 2027), as outlined in our EU-funded proposal, the Center of Excellence (CoE) has dedicated significant resources to raise awareness. Our Communication Office systematically organizes successful awareness campaigns using all available means, including social media, in full transparency. Additionally, the Biobank in collaboration with the Ethics, Legal, and Social Implications (ELSI) and Education Management, actively engages with local and regional authorities and the Human Resources departments of major enterprises. The Biobank organizes lectures, presentations, and exhibitions for young people at schools and other venues including our premises to promote biobanking and the research culture. Also, the campaigns proved effective in increasing the visibility of our website and online registration. These efforts, combined with providing engaging activities and educational material, aim to attract new volunteers to support our cause. An added value of these actions is achieving and strengthening public engagement and trust to the Biobank, preparing the next generation of citizens-scientists.

biobank.cy, since its inception, actively engages in Cyprus’s eHealth ecosystem with a focus on becoming a fully operational eHealth Biobank. Aligned with Cyprus’s 2019 eHealth law, it emphasizes citizen-centricity and interoperability. The core infrastructure includes a Biobank Information Management System (BIMS) serving as the nerve center for diverse health data, integrating clinical, sample, laboratory, genomic, and imaging data. The BIMS is designed for integration with the Cyprus’s national EHR system, using the HL7 Fast Healthcare Interoperability Resources (FHIR) standard aiming to actively contribute to primary and secondary health data use. With a vision of becoming a key player in the Cyprus National and European Health Data Space, biobank.cy designs its infrastructure to become an early adopter of initiatives like International and European Patient Summary while it already achieved integration with the BBMRI-ERIC Locator system using HL7 FHIR.

In conclusion, as with any other Biobank, the Biobank of Cyprus is creating the prospects for next-generation biomedical research. It is empowering investigations that include epidemiological and genetic determinants with the aim to identify and evaluate the contribution of factors involved in major public health burdens caused by primarily not communicable diseases.

## Acknowledgements

This work was funded by the *CY*-Biobank, which is an EU Horizon 2020 Research and Innovation Programme under Grant Agreement no. 857122, the Republic of Cyprus, and the University of Cyprus. The authors thank the research nurses, Biobank technologists, and administrative personnel who contributed to making this operation possible. The Biobank Directorship thanks all participants who voluntarily enrolled in the Biobank and made this momentum task a reality for the health benefit of all.

## Author contributions

Material preparation, participant recruitment, collection of biological material, and data collection were performed with variable contributions of all authors. Statistical analyses were performed by EML. The first draft of the manuscript was written by EML, CD, GP, AM, MK, ACK, MT, GC and SA and all authors commented on the manuscript. All authors read and approved the final manuscript.

## Availability of data and materials

The datasets used and/or analysed during the current study are available from the corresponding author, upon a reasonable request.

## DECLARATIONS

### Ethics approval and consent to participate

Ethical approval was granted from the Cyprus National Bioethics Committee, code ΕΕΒΚ/EΠ/2011/04.

### Consent for publication

Not applicable.

