## Supplementary Material 1 for "biobank.cy: The Biobank of Cyprus past, present and future"

This form provides the explanations in plain and comprehensible language regarding what is being requested from you and/or what will happen to you if you agree to join the programme:

1. All risks that may exist or any inconvenience you may incur from participating in the programme.
2. The person(s) who will have access to your information and will arise from the programme you will take part in and/or other material/data that you voluntarily provide for the programme.
3. The time period during which the Principal Investigator will have access to your information and/or material concerning you.
4. What the Principal Investigator hope to learn as a result of your participation.
5. Estimation of the benefit that can be gained for researchers and/or sponsors of this programme.
6. **You should not participate if you do not wish to, or if you have any concerns about your participation in the programme.**
7. If you decide to join, you must indicate if you have participated in any other research programmes within the last 12 months.
8. If you decide not to participate and you are a patient, your treatment will not be affected by your decision.
9. **You are free to withdraw your consent to participating in the programme at any time.**
10. If you are a patient, your decision to withdraw your consent will not have any effect on your treatment.
11. All pages of consent forms must bear your full name and signature.

Principal Investigator of the Programme you are invited to participate in

Professor Constantinos Deltas

University of Cyprus Medical School

Director, Center of Excellence in Biobanking and Biomedical Research | <https://biobank.cy/>

University of Cyprus

Programme Duration:

The Biobank has operational approval by the Cyprus National Bioethics Committee (CNBC) until 2036. After that, an approval renewal will be requested.

| Do you give consent for yourself or for someone else? |
| --- |
| If you have responded for another person, please provide details and name |

| Question | YES or NO |
| --- | --- |
| Did you fill in your consent forms personally? |  |
| Over the past 12 months, have you been involved in any other research programme? |  |
| Did you read and understand the information regarding patients and/or volunteers? |  |
| Have you had the opportunity to ask questions and discuss the Programme? |  |
| Have you been given satisfactory answers and explanations to any of your questions? |  |
| Do you understand that you can withdraw from the programme whenever you wish? |  |
| Do you understand that if you withdraw, you do not need to give any explanations for your decision? |  |
| (For patients) do you understand that, if you withdraw, there will be no impact on any treatment you get or you can get in the future? |  |
| **Do you agree to join the programme?** |  |
| With whom did you speak with? | |

Brief description of the programme (procedures and purpose).

You are invited to participate as a volunteer to the Biobank of the University of Cyprus and contribute to research programs that will be implemented and are associated with our health.

**What is a Biobank?** A Biobank is an organized collection of medical records and information that are accompanied by biological material in the form of DNA, serum, plasma, urine and other (e.g. biopsies, hair, nasopharyngeal swab etc.) with the aim to support basic or applied precision medical research. Biobanks as research infrastructures, promote the discovery of new knowledge to help each patient (personalized medicine), with the development of innovative treatments and improved processes of diagnosis and patient management.

The Biobank was created with competitive funding from the Republic of Cyprus and the European Regional Development Fund, through the Cyprus Research Promotion Foundation (Project: Strategic Infrastructure: *NEW INFRASTRUCTURE /STRATEGIC/0308/24*), and was approved by the Cyprus National Bioethics Committee in 2011 (*File No: EEBK/EΠ/2011/04*).

According to its official approval, the University of Cyprus Biobank ***specializes in kidney and other genetic diseases.***

In 2019, further funding was obtained through competition under the European Horizon 2020 program to expand this research infrastructure as part of the Center of Excellence in Biobanking and Biomedical Research.

**Main objectives of the project:**

**a.** Enhancement of the Biobank in order to support its research capacity by involving as many volunteer donors as possible, both healthy and patients.

**b.** The study of the Cypriot genome (DNA) in order to discover information that will lead to a deeper understanding of the Cypriot genetic heritage and to enhance studies for better diagnosis, prognosis and prevention of diseases.

**c.** The study of specific hereditary diseases, or diseases involving complex genetic and environmental factors, in order to acquire knowledge that will help us discover better medication for the benefit of the general public.

**How will the objectives of the program be achieved?** Your altruistic involvement is needed, as well as that of thousands of Cypriot volunteer donors, to support research studies. To do this, you need to give us access to your medical record and donate blood, and other biological material to be used in research. Your medical record may be the content of your GESY file, together with any other information you may provide to the Biobank. It is specified that access to your medical files will only be possible following your written consent, provided by signing this document.

Any research program implemented has approval by the Cyprus National Bioethics Committee. Use of your personal information will be strictly confidential and secure, in accordance with Cyprus law 125(I)/2018) for implementation of the European Data Protection Regulation **(EU) 2016/679**.

**Sending Information and Samples Abroad:** It is possible that, in the context of research programs, collaborations with researchers in other centers or universities, in Cyprus and abroad, may be undertaken in order to enhance and improve research results. Samples or coded items of your personal file may be sent to researchers abroad in the framework of such collaborations, always with SECURITY, ANONYMITY and confidentiality. By signing this consent form, you agree that the Biobank can analyse any biological material provided and use the results of these analyses for the benefit of society and the general public.

As provided by law, the processing controller is the Center of Excellence in Biobanking and Biomedical Research (biobank.cy) and the University of Cyprus. The responsibility for data protection (Data Protection Officer – DPO) lies with the legal firm KOUSIOS KORFIOTIS PAPACHARALAMBOUS (https://kkplaw.com/)

Details of what will be requested and/or what will happen to programme participants

**Registering and participating as a volunteer donor, either as a patient or as a healthy person**, is deemed useful for the success of the Biobank goals and the wider project as described above, and does not pose any risk nor will it affect the treatment you are provided or you will be provided by your doctor, if you are a patient.

You will be asked to provide personal information about yourself and your medical history and access to your medical record. You may also be asked to provide recent blood and urine test results. You will then be asked to provide some biological material as described in a different question below.

As part of your participation in the program, you will not suffer further discomfort or undergo a biopsy, if this is not medically indicated or you do not wish to. Anything that needs to be done in the context of the investigation will always be discussed with your doctor and will be performed after you have been informed and signed this consent form. The results of the research are likely to have a direct or indirect medical benefit for you, your children, or other relatives. However, it should be made clear that due to the nature of the research, exporting useful results may take many months or years or something useful may never arise for you personally or your family members.

**If you agree,** and where applicable, you will complete a questionnaire for your personal and family history and undergo a series of specialized examinations and blood tests related to your physical condition and health, such as physical measurements (height, weight, waist and hip circumference, etc.), blood pressure measurement and arterial stiffness, grip strength, spirometry, retinal image, etc. You will not incur any costs.

The above is only indicative and will be useful for your health, no matter which program you will participate in the future. If you are invited to join the Biobank under a specific research program upon your doctor's invitation, you may only undergo some of the above tests/analyses and additionally undergo some others for which your doctor will decide, ***given that the program is approved by the Cyprus National Bioethics Committee.***

Please note that it is your right to accept only part of the tests or analyses required.

**The results of haematological & biochemical tests and examinations will be shared with you during your visit to the Biobank by a healthcare professional.**

Details of the funding of the research programme

The program is funded as follows:

European Union: EUR 15 million (for 7 years, until September 2026)

Republic of Cyprus: EUR 15 million

University of Cyprus: EUR 8 million

Funding from Cypriot sources is for 15 years, until September 2034.

Details of any risks that may exist or any inconvenience that programme participants may incur

No discomfort is anticipated, beyond the collection of blood and urine or other biological material. Some people may have a small bruise at the puncture site, which will normally go away in a few days.

DNA analysis will be performed with the latest methods for determining the sequence of the “letters” in the genetic code, i.e. the nucleotides, which are the molecules making up the DNA. By using these methods, as described in this form, the program aims to the determination of the sequence of the letters of the genome in at least 500 healthy family trios. Understanding the information that lies within the DNA is a difficult and complicated process. As technology advances, by studying the DNA experts are able to uncover useful information regarding your health and other behavioural characteristics, such as your mental health, increased risk for a cardiac condition or cancer etc, as well as genetic factors protecting from some chronic conditions, such as hypertension, diabetes and dementia. For some conditions, it might be easier to come to conclusions, whereas for others great effort and discovery of new knowledge are required. Your voluntary participation will contribute towards achieving these noble goals of science and medicine. Your personal results might be ANONYMOUSLY included in local or international databases, along with the results from thousands of other volunteers across the world, for the enhancement of the world effort for combating diseases.

As with any database, an extremely small risk of a data breach cannot be completely ruled out. The University of Cyprus implements every available measure to eliminate this danger. The University of Cyprus IT Infrastructure Service, in collaboration with external advisors with expertise in data handling and protection, implements practices and mechanisms that protect against any theft or leak such as: storing data and servers in secure data centers that are under constant visual surveillance, software and virtual machine protection (Firewalls, Intrusion Detection/Prevention Systems), use of Private Network Address (IP), High Availability Architecture, use of sophisticated passwords, use of antivirus software, and implementation of data center security policies. The above measures are the best practices for data security, **to ensure data will never be acquired by non-authorised individuals and institutions, without your consent.**

Details of what information and/or what material will be collected under the programme, who will have access to it and for how long.

You will be asked for information about your demographics and your overall health, by requesting access to your medical record. From your medical record, data will be extracted about your symptoms, blood and urine tests, genetic or other tests, results and data from ultrasounds or other imaging methods, and generally anything that provides information about your health. By providing your personal information and biological material, information regarding close relatives might ensue, which will ALWAYS REMAIN CONFIDENTIAL.

You will be asked to provide biological material as follows:

a. Blood from which DNA and/or RNA, plasma, serum, cells will be isolated

b. Mouthwash or saliva for isolation of DNA and/or RNA or another biological molecule

c. Urine or hair or nasopharyngeal swab or other material

d. Genetic/protein or other material from a biopsy that you may have undergone (or will undergo in the future) for medical reasons, with your consent.

Depending on the goals of the research programs and funding in the upcoming years, your biological samples may be subjected to a series of analyses. This information will be accessible to those directly involved in the research, i.e. doctors, the project coordinator and researchers who sign a confidentiality form. The unlimited future distribution of material and data to other researchers in Cyprus or abroad will be anonymized so that the file and data cannot be linked to you.

In case of any complaint for any reason, please contact:

Marios Demetriades

Head of Research Support Service

University of Cyprus, P.O. 20537, 1678 Nicosia

Tel: (+357) 22894287 | Fax: (+357) 22895506

Access is granted until 2036, as per the Biobank operational approval. After that, an approval renewal will be requested.

If new information that directly affects your health is discovered, would you like to be informed?

YES

NO

I CANNOT MAKE A DECISION NOW. PLEASE ASK AGAIN IF NEEDED

Details of what data will be generated for you within the programme, who will have access to them and for how long.

New knowledge will emerge about the aetiology of some of the diseases that you or other people suffer, new information that will help better diagnose or even treat illnesses, knowledge that may help invent new drugs for diseases such as heart diseases, kidney diseases, cancers, etc.

Access will be for as many years as the Biobank operates legally, because it will be very useful as reference data for future research. Persons other than the research team may have access anonymously, in accordance with the approval to be obtained following an application to the CNBC.

Expected benefit for participants

If you are healthy, data that may be useful for preventative purposes may emerge. As a result, you will be able to take some measures to protect your health. If you are a patient it is likely that the genetic or other cause for your illness will be found. This may help receive better treatment in the future. This may also extend to your close relatives, such as your children. While it may be possible in the future for researchers and/or the University of Cyprus to have financial benefits from the use of research results (marketable intellectual property rights), you will not benefit financially, and you will not incur any financial burden.

Expected benefit for researchers and/or sponsors

There is no expected financial benefit to researchers or funders unless marketable copyrights arise. The main benefit for researchers is research publications in reputable journals that will give recognition and career advancement.

Details of termination or early interruption of the research programme.

No conditions for termination or early interruption of the program are anticipated

Site and duration of storage of data and/or biological samples to be collected under the programme

Your demographic and clinical data will be digitally imported and stored in the REDCap program, which is an approved and secure web application for storing the data before importing it into the Biobank database. The University of Cyprus is licensed to use this application in a secure and controlled environment.

The University of Cyprus has a dedicated suite for the storage of records and samples, which is the Biobank. All biological samples collected will be stored encoded in the Biobank in specially-locked -80^ο^C freezers located in a separate locked room, or by another freezing mechanism.

The key is only accessible to the coordinator and the person in charge of the Biobank. For the purposes of research and/or analysis of data derived from research programs, coded biological samples and anonymous results will be analysed by partners in Cyprus or abroad. The records and samples will be kept until 2036.

The present approval by the Cyprus National Bioethics Committee for the operation of the Biobank expires in 2036, after which an extension will be requested.

Description of procedures of handling data and/or biological samples of participants who withdraw from the study prior to its completion.

In order not to interfere with the credibility of the research results that may be ongoing, and to minimize the loss of scientific benefit, if you decide to withdraw, you will be asked by the project manager if you agree for the Biobank to keep all of your data and biological samples that have been collected up to that point, in full anonymity and disconnected from your identifiable folder. If you do not agree, the Biobank will delete your personal data and results and destroy the biological samples.

Full contact details and title of the person to whom participants can submit complaints or grievances regarding the programme they participate in.

Marios Demetriades

Head of Research Support Service

University of Cyprus, P.O. 20537, 1678 Nicosia

Tel: (+357) 22894287 | Fax: (+357) 22895506

Full contact details and title of the person whom participants can contact for more information or clarifications about the research programme.

Prof. Constantinos Deltas

University of Cyprus Medical School

Director, Center of Excellence in Biobanking and Biomedical Research (<https://biobank.cy/>)

1 Panepistimiou street, 2109, Nicosia

| Last name: |  | First name: |
| --- | --- | --- |
| Signature: |  | Date: |
