## Supplementary Material 2 for "biobank.cy: The Biobank of Cyprus past, present and future"

| Form Name | Field Label | Choices, Calculations, OR Slider Labels |
|---|---|---|
| Volunteer IDs | Participant ID | |
| Volunteer IDs | Identification Number | |
| Volunteer IDs | UCY number | |
| Volunteer IDs | Family number | |
| Contact and identification information | Date of visit | |
| Contact and identification information | Name | |
| Contact and identification information | Middle name | |
| Contact and identification information | Surname | |
| Contact and identification information | Telephone number | |
| Contact and identification information | E-mail | |
| Address | Street | |
| Address | Number | |
| Address | Postal code | |
| Address | City | |
| Address | Country | |
| Socio-demographic characteristics | Sex | 0, Male \| 1, Female \| 2, Prefer not to say |
| Socio-demographic characteristics | Citizenship | |
| Socio-demographic characteristics | Determination of Citizenship | |
| Socio-demographic characteristics | Date of Birth | |
| Socio-demographic characteristics | Age in years (calculated automatically) | |
| Socio-demographic characteristics | Number of Siblings (Male) | |
| Socio-demographic characteristics | Number of Siblings (Female) | |
| Socio-demographic characteristics | Marital status | 1, Single 2, Married 3, Widowed 4, Divorced 5, Cohabitation Agreement |
| Socio-demographic characteristics | Number of children (Male) | |
| Socio-demographic characteristics | Number of children (Female) | |

| | | |
|---|---|---|
| Socio-demographic characteristics | Highest level of education (successfully completed) | 1, Did not complete Elementary School\|2, Elementary School\|3, Middle School\|4, High School/Technical School\|5, Tertiary vocational school e.g. AXIK, ATI\|6, College\|7, Tertiary university (degree)\|8, Master's\|9, Ph.D. |
| Socio-demographic characteristics | Place you grew up | |
| Socio-demographic characteristics | Place of origin (town/village) of your father | |
| Socio-demographic characteristics | Place of origin (town/village) of your mother | |
| Socio-demographic characteristics | Father's Surname (for women) | |
| Socio-demographic characteristics | Are you pregnant? | 1, Yes\|2, No\|3, I don't know\|4, I don't answer |
| Socio-demographic characteristics | Gestation duration (in weeks) | |
| Socio-demographic characteristics | Nationality | 1, White - Caucasian\|2, Asian\|3, Hispanic - Latino\|4, Colored - African American\|5, Native American\|6, Ethnicity Unknown\|7, Other Ethnicity (specify) |
| Socio-demographic characteristics | Community or recognized religious minority | 1, Greek Cypriots\|5, Turkish Cypriots\|10, Maronites\|15, Latinos\|20, Armenians\|25, Muslims\|30, No answer |
| Socio-demographic characteristics | Which of the following best describes your work situation? | 0, Private employee\|1, Public employee (permanent or indefinite)\|2, Public employee (temporary or fixed-term)\|3, Self-employed with employees\|4, Self-employed without employees\|5, Retired\|6, Housewife\|7, Farmer\| 8, Unemployed\|9, Other |
| Socio-demographic characteristics | Please describe your employment status | |
| Socio-demographic characteristics | Has your employment been full-time or part-time in the last year? | 1, Full\|2, Part\|3, Not working |

| | | |
|---|---|---|
| Socio-demographic characteristics | Please indicate the occupation you have been in for most years | |
| Socio-demographic characteristics | In which of the following categories does your household's net monthly income (i.e. if you add the net monthly income of all members of your household) in euros fall into | 1, Up to 900\|2, 901-1200\|3, 1201-1500\|4, 1501-1900\|5, 1901-2300\|6, 2300-2800\|7, 2801-3500\|8, 3501-4000\|9, 4001 - 5500\|10, 5501 and above\|11, Don't know/not sure/don't remember\|12, Don't answer |
| Socio-demographic characteristics | Are you facing any form of financial difficulty in your daily needs/bill payments? | 1, Yes\|2, No |
| Quality of Life | Please mark on the scale below the number that best represents your state of health, where 100 is the best state you can imagine and 0 is the worst | |
| Quality of Life | Do you smoke regularly (>1 cigarette every day)? | 1, Yes\|2, No\|3, In the past (>1 timeout) |
| Quality of Life | At what age did you stop smoking? | |
| Quality of Life | At what age did you start smoking? | |
| Quality of Life | How many cigarettes do you smoke a day? | |
| Quality of Life | How many packs of cigarettes do you smoke a week? | |
| Quality of Life | How often do you drink alcoholic beverages? | 1, Never\|2, Monthly or less\|3, 2-4 times a month\|4, 2-3 times a week\|5, 4 or more times a week |
| Quality of Life | Do you have a long-term illness or chronic health problem? | 1, Yes\|2, No\|3, I don't know/I'm not sure/ I don't remember\|4, I don't answer |
| Quality of Life | During the past 6 months or more, have you restricted your usual activities because of a health problem, and if so, to what extent? | 1, Yes, to a significant extent\|2, Yes, to some extent\|3, No limitation\|4, Don't know/not sure/don't remember\|5, Don't answer |
| Vital Signs | Height in centimeters (cm) | |

| | | |
|---|---|---|
| Vital Signs | Weight in kilograms (Kg) | |
| Vital Signs | Body Surface Area - BSA | |
| Vital Signs | Body Mass Index - BMI | |
| Vital Signs | Blood pressure - Systolic mmHg | |
| Vital Signs | Blood pressure - Diastolic mmHg | |
| Vital Signs | Waist circumference (cm) | |
| Physical Activity - IPAQ | During the past 7 days, how many days did you do any vigorous physical activity, such as digging, vigorous weight training, running on an incline, brisk running, aerobics, brisk cycling, brisk swimming, singles tennis, playing on a field (soccer, basketball-basketball, volleyball, etc.? | 1, 0 / 2, 1 / 3, 2 / 4, 3 / 5, 4 / 6, 5 / 7, 6 / 8, 7 |
| Physical Activity - IPAQ | On days you did some vigorous physical activity, how much time did you usually spend? (in minutes of the hour) | 1, 10 / 2, 20 / 3, 30 / 4, 40 / 5, 50 / 6, 60 / 7, 70 / 8, 80 / 9, 90 / 10, Don't know / Not sure / 11, Other |
| Physical Activity - IPAQ | Please specify the time in minutes of the hour | |
| Physical Activity - IPAQ | During the past 7 days, how many days did you do any moderate physical activity, such as lifting and carrying light weights (less than 10 kg), general house cleaning, gentle rhythmic body exercises, leisurely cycling at a slow speed, leisurely swimming? Please do not include walking. | 1, 0 / 2, 1 / 3, 2 / 4, 3 / 5, 4 / 6, 5 / 7, 6 / 8, 7 |
| Physical Activity - IPAQ | On days you did some moderate physical activity, how long did you usually spend (in minutes per hour)? | 1, 10 / 2, 20 / 3, 30 / 4, 40 / 5, 50 / 6, 60 / 7, 70 / 8, 80 / 9, 90 / 10, Don't know / Not sure / 11, Other |
| Physical Activity - IPAQ | Please specify the time in minutes of the hour | |

| | | |
|---|---|---|
| Physical Activity - IPAQ | During the past 7 days, how many days did you walk for more than 10 minutes in a row? | 1, 0 \| 2, 1 \| 3, 2 \| 4, 3 \| 5, 4 \| 6, 5 \| 7, 6 \| 8, 7 |
| Physical Activity - IPAQ | On days you walked for more than 10 minutes straight, how much time did you spend walking (in minutes per hour)? | 1, 10 \| 2, 20 \| 3, 30 \| 4, 40 \| 5, 50 \| 6, 60 \| 7, 70 \| 8, 80 \| 9, 90 \| 10, Don't know / Not sure \| 11, Other |
| Physical Activity - IPAQ | Please specify the time in minutes of the hour | |
| Physical Activity - IPAQ | During the past 7 days, how much time did you spend sitting on a typical day? This time may include time spent sitting at home, in the office, in the car, when reading, when with friends, resting in an armchair or watching TV, but does not include sleep (in hours per day). | 1, 0 \| 2, 1 \| 3, 2 \| 4, 3 \| 5, 4 \| 6, 5 \| 7, 6 \| 8, 7 \| 9, 8 \| 10, 9 \| 11, 10 \| 12, 11 \| 13, 12 \| 14, 13 \| 15, 14 \| 16, 15 \| 17, Don't know / Not sure \| 18, Other |
| Physical Activity - IPAQ | Please specify the time in hours per day | |
| Physical Activity - IPAQ | a) at work (work) | 1, Strong \| 2, Moderate \| 3, Low \| 4, Don't know |
| Physical Activity - IPAQ | b) in transportation | 1, Strong \| 2, Moderate \| 3, Low \| 4, Don't know |
| Physical Activity - IPAQ | c) chores in and around the home (including housekeeping, gardening, general repairs or looking after the family) | 1, Strong \| 2, Moderate \| 3, Low \| 4, Don't know |
| Physical Activity - IPAQ | d) for entertainment, sports and leisure activities | 1, Strong \| 2, Moderate \| 3, Low \| 4, Don't know |
| Diet - MedDietScore | Frequency of consumption of whole grains (e.g. bread, pasta, rice, etc.) (servings/week) | 1, Never \| 2, 1-6 \| 3, 7-12 \| 4, 13-18 \| 5, 19-31 \| 6, >32 |
| Diet - MedDietScore | Frequency of potato consumption (servings/week) | 1, Never \| 2, 1-4 \| 3, 5-8 \| 4, 9-12 \| 5, 13-18 \| 6, >18 |
| Diet - MedDietScore | Frequency of fruit and juice consumption (portions/week) | 1, Never \| 2, 1-4 \| 3, 5-8 \| 4, 9-15 \| 5, 16-21 \| 6, >22 |

<table>
<tr><td>Diet - MedDietScore</td><td>Frequency of consumption of vegetables and salads (portions/week)</td><td>1, Never|2, 1-6|3, 7-12|4, 13-20|5, 21-32|6, >33</td></tr>
<tr><td>Diet - MedDietScore</td><td>Frequency of consumption of legumes (servings/week)</td><td>1, Never|2, < 1|3, 1-2|4, 3-4|5, 5-6|6, >6</td></tr>
<tr><td>Diet - MedDietScore</td><td>Frequency of fish and soup consumption (portions/week)</td><td>1, Never|2, < 1|3, 1-2|4, 3-4|5, 5-6|6, >6</td></tr>
<tr><td>Diet - MedDietScore</td><td>Frequency of consumption of red meat and its products (portions/week)</td><td>1, <=1|2, 2-3|3, 4-5|4, 6-7|5, 8-10|6, >10</td></tr>
<tr><td>Diet - MedDietScore</td><td>Frequency of poultry consumption (servings/week)</td><td>1, <=3|2, 4-5|3, 5-6|4, 7-8|5, 9-10|6, >10</td></tr>
<tr><td>Diet - MedDietScore</td><td>Frequency of full-fat dairy consumption (servings/week)</td><td>1, <=10|2, 11-15|3, 16-20|4, 21-28|5, 29-30|6, >30</td></tr>
<tr><td>Diet - MedDietScore</td><td>Frequency of olive oil consumption in daily cooking (portions/week)</td><td>1, Never|2, Rarely|3, < 1|4, 1-3|5, 3-5|6, daily</td></tr>
<tr><td>Diet - MedDietScore</td><td>Frequency of consumption of alcoholic beverages (ml/day, 100 ml = 1 glass 12%)</td><td>1, < 300|2, 300|3, 400|4, 500|5, 600|6, >700 or 0</td></tr>
<tr><td>Diet - MedDietScore</td><td>MedDietScore (c) (Calculated automatically)</td><td></td></tr>
<tr><td>Diet</td><td>In the last month you systematically took:</td><td>1, Vitamins|2, Protein|3, Creatine|4, Other</td></tr>
<tr><td>Diet</td><td>Please specify nutritional supplements</td><td></td></tr>
<tr><td>Sleep Assessment - GSAQ</td><td>Did you have difficulty falling asleep, staying asleep, or felt insufficiently rested in the morning?</td><td>1, Never|2, Sometimes|3, Usually|4, Always</td></tr>
<tr><td>Sleep Assessment - GSAQ</td><td>Have you fallen asleep involuntarily or have to struggle to stay awake during the day?</td><td>1, Never|2, Sometimes|3, Usually|4, Always</td></tr>
<tr><td>Sleep Assessment - GSAQ</td><td>Are sleep difficulties or daytime sleepiness affecting your daily activities?</td><td>1, Never|2, Sometimes|3, Usually|4, Always</td></tr>
<tr><td>Sleep Assessment - GSAQ</td><td>Does work or other activities prevent you from getting enough sleep?</td><td>1, Never|2, Sometimes|3, Usually|4, Always</td></tr>
<tr><td>Sleep Assessment - GSAQ</td><td>Do you fart loudly?</td><td>1, Never|2, Sometimes|3, Usually|4, Always</td></tr>
</table>

| | | |
|---|---|---|
| Sleep Assessment - GSAQ | Have you held your breath, had pauses in breathing, or stopped breathing in your sleep? | 1, Never\|2, Sometimes\|3, Usually\|4, Always |
| Sleep Assessment - GSAQ | Have you had restless legs or feel like you're dragging them at night? Does the sensation go away if you move your legs? | 1, Never\|2, Sometimes\|3, Usually\|4, Always |
| Sleep Assessment - GSAQ | Have you had repetitive rhythmic leg jerks or leg spasms during your sleep? | 1, Never\|2, Sometimes\|3, Usually\|4, Always |
| Sleep Assessment - GSAQ | Have you had nightmares, or screamed, walked, punched or kicked in your sleep? | 1, Never\|2, Sometimes\|3, Usually\|4, Always |
| Sleep Assessment - GSAQ | Were the following things disturbing your sleep: pain, other physical symptoms, worries, medications, or other (specify)? | 1, Never\|2, Sometimes\|3, Usually\|4, Always |
| Sleep Assessment - GSAQ | Did you feel sad or anxious? | 1, Never\|2, Sometimes\|3, Usually\|4, Always |
| Clinical part | Last Visit to the Doctor | |
| Clinical part | Personal Doctor's name | |
| Clinical part | Personal Doctor's phone number | |
| Clinical part | Name of Nephrologist | |
| Clinical part | Nephrologist phone number | |
| Clinical part | Age of first menstruation | |
| Clinical part | You used contraceptives | 1, Yes\|2, No |
| Clinical part | For how many years? | |
| Clinical part | Menopause; | 1, Yes\|2, No |
| Clinical part | Age of menopause | |
| Clinical part - medical conditions | Hypertension (high blood pressure) | 1, Yes\|2, No\|3, Don't know - Not sure\|4, Don't answer |
| Clinical part - medical conditions | Asthma (including allergic) | 1, Yes\|2, No\|3, Don't know - Not sure\|4, Don't answer |
| Clinical part - medical conditions | Chronic bronchitis, chronic obstructive pulmonary disease, emphysema | 1, Yes\|2, No\|3, Don't know - Not sure\|4, Don't answer |
| Clinical part - medical conditions | Myocardial infarction (heart attack) or chronic effects of an old heart attack | 1, Yes\|2, No\|3, Don't know - Not sure\|4, Don't answer |

<table>
<tr><td>Clinical part - medical conditions</td><td>Stroke (bleeding in the brain, clot in the brain) or chronic effects of an old stroke</td><td>1, Yes | 2, No | 3, Don't know - Not sure | 4, Don't answer</td></tr>
<tr><td>Clinical part - medical conditions</td><td>Joint disease (excluding arthritis)</td><td>1, Yes | 2, No | 3, Don't know - Not sure | 4, Don't answer</td></tr>
<tr><td>Clinical part - medical conditions</td><td>Rheumatoid arthritis</td><td>1, Yes | 2, No | 3, Don't know - Not sure | 4, Don't answer</td></tr>
<tr><td>Clinical part - medical conditions</td><td>Lupus erythematosus</td><td>1, Yes | 2, No | 3, Don't know - Not sure | 4, Don't answer</td></tr>
<tr><td>Clinical part - medical conditions</td><td>Osteoarthritis</td><td>1, Yes | 2, No | 3, Don't know - Not sure | 4, Don't answer</td></tr>
<tr><td>Clinical part - medical conditions</td><td>Osteoporosis</td><td>1, Yes | 2, No | 3, Don't know - Not sure | 4, Don't answer</td></tr>
<tr><td>Clinical part - medical conditions</td><td>Low back pain or other chronic back problems (back pain - disc disease)</td><td>1, Yes | 2, No | 3, Don't know - Not sure | 4, Don't answer</td></tr>
<tr><td>Clinical part - medical conditions</td><td>Neck disease or other chronic neck problems</td><td>1, Yes | 2, No | 3, Don't know - Not sure | 4, Don't answer</td></tr>
<tr><td>Clinical part - medical conditions</td><td>Type I diabetes</td><td>1, Yes | 2, No | 3, Don't know - Not sure | 4, Don't answer</td></tr>
<tr><td>Clinical part - medical conditions</td><td>Type II diabetes</td><td>1, Yes | 2, No | 3, Don't know - Not sure | 4, Don't answer</td></tr>
<tr><td>Clinical part - medical conditions</td><td>Allergy (e.g. rhinitis, dermatitis, food allergy, etc. - excluding allergic asthma)</td><td>1, Yes | 2, No | 3, Don't know - Not sure | 4, Don't answer</td></tr>
<tr><td>Clinical part - medical conditions</td><td>Cirrhosis of the liver</td><td>1, Yes | 2, No | 3, Don't know - Not sure | 4, Don't answer</td></tr>
<tr><td>Clinical part - medical conditions</td><td>Kidney diseases</td><td>1, Yes | 2, No | 3, Don't know - Not sure | 4, Don't answer</td></tr>
<tr><td>Clinical part - medical conditions</td><td>Cancer (malignancy, leukemia, lymphoma)<br>If yes, please specify e.g. breast, neck, lung, etc</td><td>1, Yes | 2, No | 3, Don't know - Not sure | 4, Don't answer</td></tr>
<tr><td>Clinical part - medical conditions</td><td>Depression</td><td>1, Yes | 2, No | 3, Don't know - Not sure | 4, Don't answer</td></tr>
<tr><td>Clinical part - medical conditions</td><td>Autism</td><td>1, Yes | 2, No | 3, Don't know - Not sure | 4, Don't answer</td></tr>
</table>

| | | |
|---|---|---|
| Clinical part - medical conditions | Alzheimers | 1, Yes\|2, No\|3, Don't know - Not sure\|4, Don't answer |
| Clinical part - medical conditions | Have you been diagnosed with any other medical condition besides the above? Please specify | |
| Clinical part | Please specify the cancer | |
| Clinical part | Are you taking any medication from your doctor? | 1, Yes\|2, No |
| Clinical part | Please specify medication (SNOMED CT) | |
| Clinical part | At what age or year was Diabetes Mellitus first diagnosed? | |
| Clinical part | Do you take insulin? | 1, Yes\|2, No |
| Clinical part | At what age did you start taking insulin? | |
| Clinical part | Have you ever had your urine dipstick checked for microalbuminuria, | 1, Yes\|2, No |
| Clinical part | Have you had anything out of the ordinary? | |
| Clinical part | Have you had a kidney biopsy? | 1, Yes\|2, No |
| Clinical part | When did you have a kidney biopsy? | |
| Clinical part | What were the kidney biopsy findings? | |
| Clinical part | Define heart disease | |
| Clinical part | Have you ever had a cardiovascular system checkup (Electrocardiogram, stress test, echocardiogram or other test)? | 1, Yes\|2, No |
| Clinical part | What were the results of the audit? | |
| Clinical part | Have you ever had a coronary angiogram? | 1, Yes\|2, No |
| Clinical part | What were the results of the audit? | |
| Clinical part | Other abnormal findings or tests from the cardiovascular system? | |
| Clinical part | What is the diagnosis? (ICD10) | |
| Clinical part | Have you ever had a liver ultrasound? | 1, Yes\|2, No |
| Clinical part | What were the findings? | |

| | | |
|---|---|---|
| Clinical part | Have you ever had a liver biopsy? | 1, Yes\|2, No |
| Clinical part | Liver biopsy date? | |
| Clinical part | What were the liver biopsy findings? | |
| Clinical part | Have you had any special tests (blood/urine tests) for liver dysfunction? | 1, Yes\|2, No |
| Clinical part | What tests and what were the findings? | |
| Clinical part | Please fill in any observations or comments | |
| Clinical part | You have been diagnosed with any medical condition, e.g. Type I/Type II Diabetes, Asthma, Hypertension or other? | 1, Yes\|2, No |
| Clinical part | Please specify the condition | |
| Clinical part | Are you taking any medication from your doctor? | 1, Yes\|2, No |
| Clinical part | Please specify the medication | |
| Clinical part | Is the patient healthy? Can it be included in the Healthy Control Cohort? | 1, Yes\|2, No |
| Clinical part | Why can't it be included in the healthy? | |
| family_health_history | How many of your grandparents lived to be over 70? | |
| family_health_history | How many of your grandparents lived to be over 100 years old? | |
| family_health_history | Are there kidney function problems (kidney failure) in your family? | |
| family_health_history | Have you or anyone in your family ever had a heart attack, heart attack, angina, stroke or other heart disease? | 1, Yes\|2, No |
| family_health_history | Have you or anyone in your family ever had liver disease or liver dysfunction (eg, hepatitis, cirrhosis, ascites, liver malignancy, fatty liver, hemochromatosis, etc.)? | 1, Yes\|2, No |

| | | |
|---|---|---|
| family_health_history | Do you know if there is any of the following medical conditions and/or documented diagnoses in your family that affect more than 2 people? | 1, Diabetes Mellitus Type 1 \| 2, Diabetes Mellitus Type 2 \| 3, Asthma \| 4, Neurological conditions/diagnoses \| 5, Coronary Heart Disease \| 6, Kidney Disease or Kidney Dysfunction \| 7, Hypertension \| 8, Other |
| family_health_history | Is there a diagnosis of neoplasia (cancer) in your family? | 1, Yes \| 2, No |
| family_health_history | Please specify the type of cancer | |
| Biochemistry hematology | Glucose | |
| Biochemistry hematology | Uric Acid | |
| Biochemistry hematology | Glycosylated Haemoglobin A1c | |
| Biochemistry hematology | Creatinine | |
| Biochemistry hematology | Total Cholesterol | |
| Biochemistry hematology | HDL Cholesterol | |
| Biochemistry hematology | Triglycerides | |
| Biochemistry hematology | LDL Cholesterol | |
| Biochemistry hematology | Glucose, fasting | |
| Biochemistry hematology | Red Blood Cells (RBC) | |
| Biochemistry hematology | Hematocrit | |
| Biochemistry hematology | White Blood Cells (WBC) | |
| Biochemistry hematology | Neutrophils | |
| Biochemistry hematology | Neutrophils % | |
| Biochemistry hematology | Lymphocytes | |
| Biochemistry hematology | Lymphocytes % | |
| Biochemistry hematology | Monocyte | |
| Biochemistry hematology | Monocyte % | |
| Biochemistry hematology | Eosinophils | |
| Biochemistry hematology | Eosinophils % | |
| Biochemistry hematology | Basophils | |
| Biochemistry hematology | Basophils % | |
| Biochemistry hematology | Large Unstained Cells (LUC) | |
| Biochemistry hematology | Large Unstained Cells (LUC) % | |
| Biochemistry hematology | Serum Creatinine | |

| | |
|---|---|
| urine_biochemistry | Urine microalbumin (Microalbumin) (Urine) |
| urine_biochemistry | Urine Creatinine (Urine) |
| urine_biochemistry | Microalbumin - Creatinine Ratio (ACR) (Urine) |
| urine_biochemistry | Urine Proteins (Urine) |
| urine_biochemistry | Proteins Creatinine Ratio PCR (Urine) |
| urine_biochemistry | Clarity |
| urine_biochemistry | Color |
| urine_biochemistry | Reaction (PH) |
| urine_biochemistry | Specific Gravity |
| urine_biochemistry | Albumin |
| urine_biochemistry | Glucose |
| urine_biochemistry | Acetone - Ketones |
| urine_biochemistry | Bilirubin |
| urine_biochemistry | Urobilinogen |
| urine_biochemistry | Nitrates |
| urine_biochemistry | Erythrocytes - Red Blood Cells |
| urine_biochemistry | Leukocytes - White Blood Cells |
| urine_biochemistry | Leukocytes |
| urine_biochemistry | Erythrocytes |
| urine_biochemistry | Squamous Epithelia |
| urine_biochemistry | Casts |
| urine_biochemistry | Crystals |
| urine_biochemistry | Microorganisms |
| urine_biochemistry | Fungi |
| urine_biochemistry | eGFR MDRD |
| urine_biochemistry | MDRD based CKD Stage (GFR, mL/min/1.73 m2) |
| urine_biochemistry | eGFR CKD-EPI |
| urine_biochemistry | CKD-EPI based CKD Stage (GFR, mL/min/1.73 m2) |
